# Individualized Interactomes from Pulmonary Arterial Hypertension Cell Biopsies Predict Therapeutic Response

**DOI:** 10.1101/2025.11.13.25340200

**Authors:** Rui-Sheng Wang, Navneet Singh, Henri Wathieu, Grayson Baird, Katherine Cox-Flaherty, James R. Klinger, Christopher J. Mullin, Mandy Pereira, Mary Whittenhall, Elizabeth O. Harrington, Bradley A. Maron, Corey E. Ventetuolo

## Abstract

Pulmonary arterial hypertension (PAH) is characterized by molecular heterogeneity and variable pharmacotherapeutic responses. We used an *in silico* clinical trial design to test whether transcriptomic data from pulmonary artery endothelial cell biopsies collected during right heart catheterization could be used to build individualized protein-protein interactomes and inform treatment response. Twenty-five PAH participants (56 [range: 30-84] yr; 91% female; 81%, white) and three controls contributed 32 and three cell biopsy specimens, respectively. Patient-specific interactomes varied widely in topology and complexity. All 32 individualized PAH interactomes were enriched significantly with key PAH endophenotypes genes such as hypoxia, oxidant stress, and apoptosis. Concordance between the molecular targets of a prescribed PAH pharmacotherapy class and the corresponding individualized interactome topology was associated with improvement in multiple PAH metrics, including a reduction in brain natriuretic peptide levels at 6 months (β -627.6 pg/mL, [95% CI -986.4, -268.8]; p = 0.003) and a decrease from high to intermediate REVEAL 2.0 Risk scores at 6 months (β -1.3 units, [95% CI -2.6, -0.04]; p=0.043) that persisted at 12 months (β -1.7, [95% CI -3.4, -0.1]; p=0.059). Integration of transcriptomics acquired at point-of-care with network medicine may individualize treatment selection and improve clinically relevant endpoints in PAH.

## Introduction

Pulmonary arterial hypertension (PAH) is a highly morbid cardiopulmonary disease characterized by substantial molecular and clinical heterogeneity which drive variable treatment-response patterns observed in clinical trials and real-world practice.^1–4^ More than 16 medications spanning multiple drug classes are now approved for PAH, and there is increasing emphasis on goal-oriented, combination therapy.^5^ Utilizing patient-specific pathogenetic information to inform drug development, clinical trial design, and treatment selection in clinical practice may improve efficacy and effectiveness of PAH drugs, respectively, but such strategies are lacking. Progress achieving this goal has been hindered by challenges inherent to sampling affected tissue in PAH and the availability of analytical methods that align individual patient-level pathobiological data to treatment.

A hallmark PAH endophenotype is pulmonary artery endothelial cell (PAEC) dysfunction; *ex vivo* studies of PAECs from PAH patients validate numerous disease-specific pharmacotherapeutic targets experimentally, but these samples derive largely from end-stage patients at the time of lung explant^6–8^. We and others have described a minimally invasive “cell biopsy” approach in which PAECs can be collected from the tips of pulmonary artery catheters and propagated *in vitro*^9–12^. Thus, right heart catheterization (RHC) with associated PAEC biopsy provides a direct window into the lung vasculature by sampling a fundamental tissue compartment involved in PAH pathogenesis during a procedure that is central to the diagnosis and longitudinal management of patients.

We recently innovated a network medicine method using transcriptomic data to generate individualized interactomes that emphasize functionally relevant protein-protein interactions (PPIs).^13^ This approach captured patient-specific molecular features (that are otherwise marginalized when using reductionist analytical methods), which could then be used to predict clinical traits in cardiomyopathy patients. In the current study, we established a transcriptome-interactome-systems pharmacology bioinformatics pipeline using PAECs collected at point-of-care and implemented this retrospectively using a blinded clinical trial design *in silico*. We hypothesized that the proportion of therapeutic targets of the prescribed PAH therapy found in each cell-biopsy individualized interactome would be associated with improved clinical outcomes in PAH. Findings from this study are positioned to advance precision medicine by linking patient level molecular data to treatment selection and, ultimately, therapeutic efficacy and clinical effectiveness.

## Methods

### Overall Study Design

The overall study design is presented in **Figure 1**. This was a retrospective analysis of patients undergoing RHC for the routine clinical assessment of dyspnea or PAH monitoring at Rhode Island Hospital (Brown University Health) between 2017-2022. All patients were treated based on usual standard-of-care (informed by RHC and other results) and at the discretion of their managing PAH clinician. Clinical data were paired retrospectively with data from patient-specific PPI interactomes assembled from individual PAEC biopsies performed at the time of RHC. A systems pharmacology approach was then used to determine the molecular targets of the exposure, which in this study were FDA approved PAH therapeutics (or non-PAH drugs as comparators) prescribed at the time of, or following, RHC. Our goal was to determine if the proportion of therapeutic targets in the individual-specific network was predictive of validated clinical outcome measures assessed as part of routine clinical practice.

**Figure 1.**
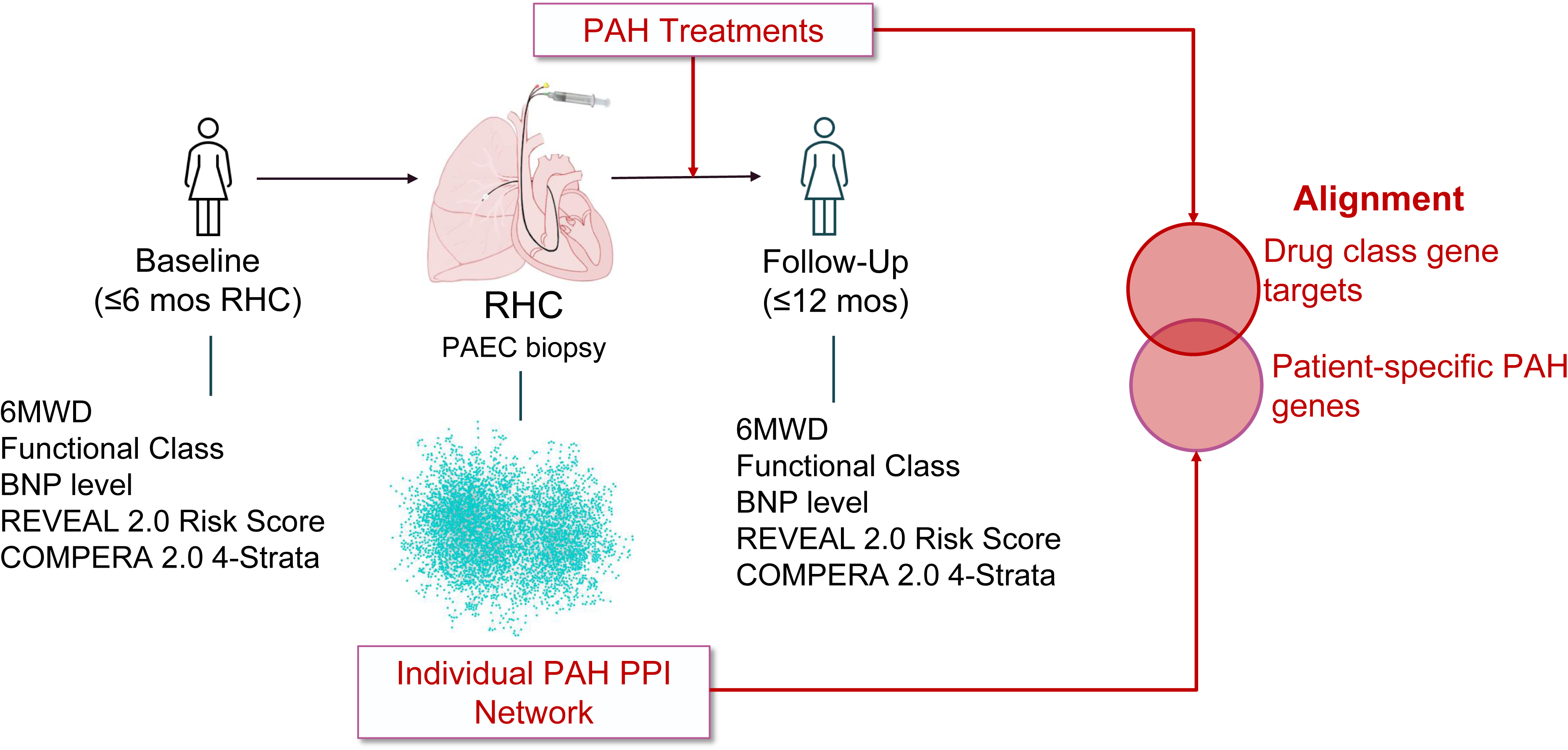
Study schematic for the *in silico* clinical trial. Clinical data was collected within six months before RHC and up to 12 months following RHC. PAH treatments were collected at the time of or after completion of the RHC, during which time a cell biopsy was obtained. Following completion of RNA sequencing, an individual PAH PPI network was constructed. The study exposure was alignment between drug class gene targets and patient specific PAH genes identified in the individual PPI. A similar approach was taken for active and inactive comparators. RHC=right heart catheterization; 6MWD=six-minute walk distance; BNP=brain natriuretic peptide; REVEAL=Registry to Evaluate Early and Long-Term PAH Disease Management; COMPERA=Comparative, Prospective Registry of Newly Initiated Therapies for PH; PAEC=pulmonary artery endothelial cell; PAH=pulmonary arterial hypertension; PPI=protein-protein interaction.

### Clinical Study Design and Data Collection

Participants were included in this study if they had a clinical diagnosis of PAH using the World Symposium on Pulmonary Hypertension criteria consistent with this diagnosis^14^ (confirmed post hoc by N.S.), had a successful PAEC biopsy from RHC^9, 10^, and had ≥ 1 follow-up clinical assessment after RHC. Baseline clinical data were collected for participants ≤6 months before the date of the study RHC. Post-RHC clinical assessments (e.g., 6-minute walk distance [6MWD], risk scores) that occurred up to 12 months after initiation of PAH therapies were included as study end points.

#### Clinical end points

We selected widely employed and established clinical metrics in PAH as our study end points. The 6MWD is the primary end point for almost all Phase III trials and the basis for drug approval in PAH. The minimal clinically important distance for 6MWD is 30-35 m^15^. Brain-natriuretic peptide (BNP) and N-terminal pro-BNP (NT-proBNP) levels are non-specific markers of right ventricular stretch that are prognostically meaningful in PAH. The cut-off values for BNP of <50, 50-199, 200-800, and >800 ng/L are ascribed to escalating risk in various multiparameter prognostic models^5^. Several of these risk assessment scores have been developed for the prognosis and monitoring of PAH and are recommended at diagnosis and frequently thereafter; World Health Organization functional class (WHO-FC), 6MWD, and BNP/NT-proBNP are the most informative components in all scores. Functional class is one of the most established clinical parameters evaluated in PAH and until recently was the basis of therapeutic escalation.

The United States Registry to Evaluate Early and Long-Term PAH Disease Management (REVEAL) 2.0 risk calculator has been validated as a prognostic score for predicting 1-year survival^16^. REVEAL 2.0 incorporates 13 total items including 6MWD, functional class and BNP as well as PAH subgroup, demographic information, hospitalizations, echocardiographic, vital sign, hemodynamic, pulmonary and renal function data. Seven components are required to generate a score and missing items are assigned “0”s otherwise; risk scores of 0-6, 7-8, and ≥9 correspond to low, intermediate and high-risk for mortality. The Comparative, Prospective Registry of Newly Initiated Therapies for PH (COMPERA) 2.0 four-strata risk assessment (1=low; 2=intermediate-low; 3=intermediate high; 4=high) for mortality incorporates functional class, 6MWD, and BNP/NT-proBNP. COMPERA 2.0 four-strata risk assessment is recommended to follow PAH patients longitudinally and is the basis for treatment escalation in the most recent consensus guidelines^5,17^. Both the clinical data extraction (N.S., H.W., K.C.F., C.E.V.) and bioinformatic team (R-S.W., B.A.M.) members worked independently such that PAH treatments and clinical metrics as well as PPI network results were blinded, respectively. The study was approved by the IRB at Brown University Health (IRB#001218).

### Cell Biopsy, RNA Sequencing, and Protein-Protein Interactome Derivation

Details of the isolation and propagation of PAECs from RHC, RNA sequencing and analysis and construction of the individualized PPI networks are included in the online supplement.

### Systems Pharmacology

We considered 10 Food and Drug Administration-approved drugs for PAH, five calcium channel blockers and vasodilators that were previously included in the PAH module and added the investigational drugs DHEA (dehydroepiandrosterone) and sotatercept, given participants in our center were enrolled in these trials^18–20^ (**Table S1**). We extracted the targets of these drugs from DrugBank (**Table S2**) and calculated whether the targets of the drugs were in the individualized PPI networks; if so, the number of targets (among all the targets of the drugs) were quantitated^21^. Target-interactome alignment, the exposure for this study, was defined as the proportion of protein targets for a given PAH drug class (e.g., phosphodiesterase-5 inhibitors) in a cell biopsy PPI network out of all the possible protein targets in that same drug class (e.g., phosphodiesterase-5 inhibitors). In other words, target-interactome alignment was the proportion of drug class targets present in the cell-biopsy-derived individualized PPI interactome for the therapy administered to a corresponding patient.

#### Control Comparators

To simulate a placebo-controlled condition in this *in silico* clinical study, we next chose concomitant medications that are prescribed frequently in PAH but are not considered PAH-targeted therapies nor are they FDA-approved for the treatment of PAH. We assessed if the proportion of protein targets for those concomitant drug classes in the sample-specific network was associated with the same clinical end points described above. Specifically, the protein targets of individual diuretic, antidepressant, and anxiolytic medications were compiled from DrugBank (Tables S1, S2). The degree of overlap of the drug targets of these ‘placebo’ drugs to the PAH disease module was calculated to make sure that the ‘placebo’ drugs were not significantly close to the PAH disease module (in essence, the degree to which inactive comparators could be considered active comparators)^21^.

We separated diuretics, antidepressants and anxiolytics into two groups: those with PAH module overlap and those without. Spironolactone was the only diuretic with overlap among the prescribed diuretics. Based on mechanism of action and correlative experimental data, several antidepressants targeting serotonin signaling^22–24^ (venlafaxine, fluoxetine, quetiapine, escitalopram, desvenlafaxine, mirtazapine, citalopram) had potential overlap with the PAH module and were, therefore, analyzed separately (i.e., active comparators; grouped as Serotonin Psychotropics) from those that did not (bupropion and benzodiazepines including clonazepam, alprazolam, lorazepam) (i.e., inactive comparators; grouped as Non-Serotonin Psychotropics).

### Statistical Analysis

Summary data are presented as median (range) and categorical data as frequency (percentage). Outcomes were modeled as a function of the proportion of protein targets in the sample-specific networks by drug class. Generalized mixed modeling with sandwich estimation assuming normal distributions with the GLIMMIX procedure was employed to account for repeated samples from some participants (25 subjects had a total of 7 biological replicates; 1 subject with 5 balloon tips, 3 subjects with 2 balloon tips each) and time. Interval estimates were calculated for 95% confidence and alpha was established at the 0.05 level. Analyses were conducting using SAS Software 9.4 (SAS Inc. Cary, NC) (G.B.)

## Results

### Cohort Characteristics

The final study cohort included 25 PAH participants and 3 controls (individuals who underwent RHC without pulmonary hypertension) contributing 32 and 3 cell biopsy specimens, respectively. The average age of PAH participants was 56 (range 30 – 84) years old and most were female (91%) and white (81%). Two participants (6%) were on no PAH-specific treatment at the time of RHC. Most participants were WHO-FC II at the time of RHC with a median 6MWD of 375 (range 150 – 575) m and a median REVEAL 2.0 Risk score of 10 (range 4 – 12). **Table 1** includes characteristics of the participants at the time of RHC, and **Table S3** categorizes patient subgroups according to PAH treatment received post-RHC.

**Table 1.**
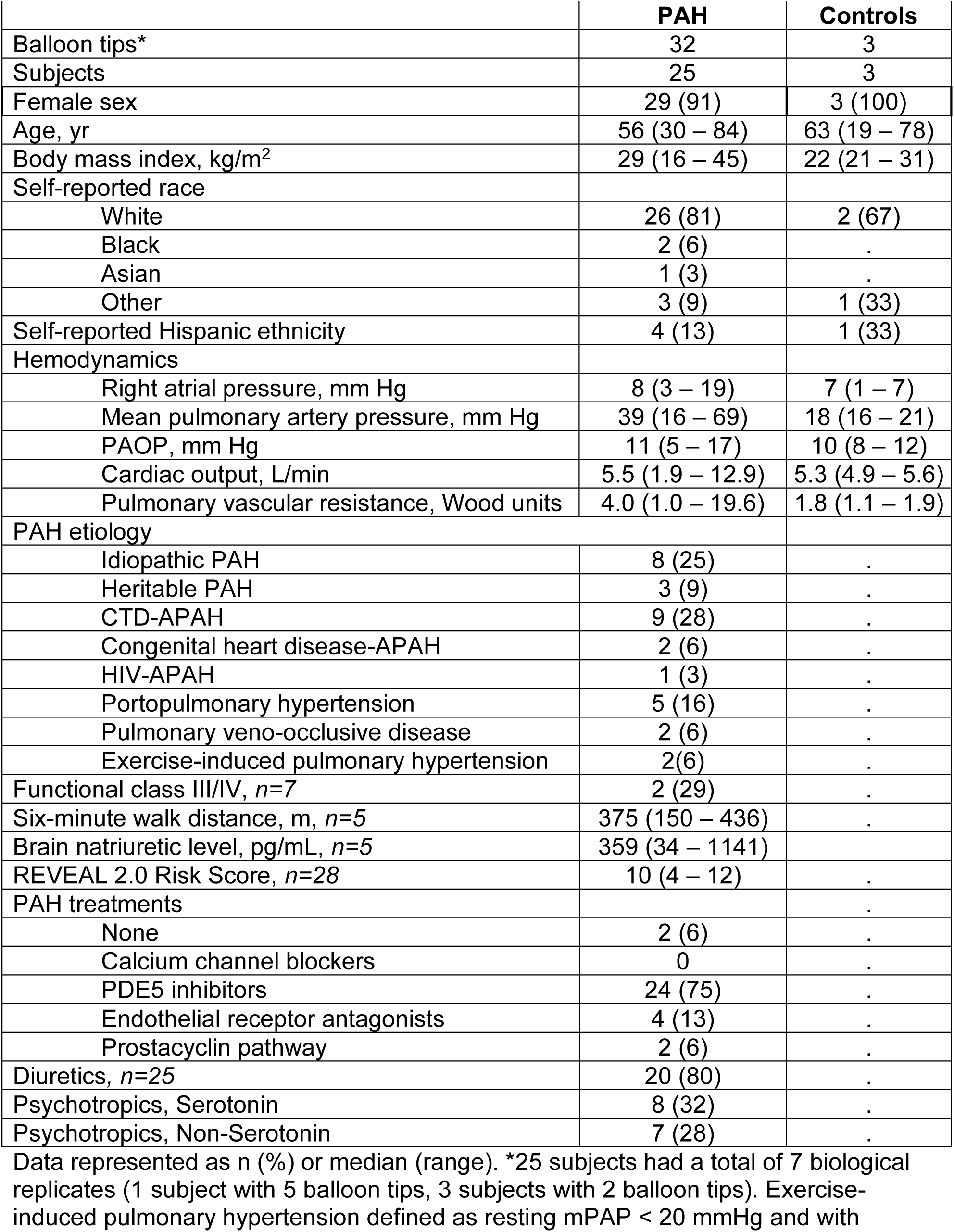
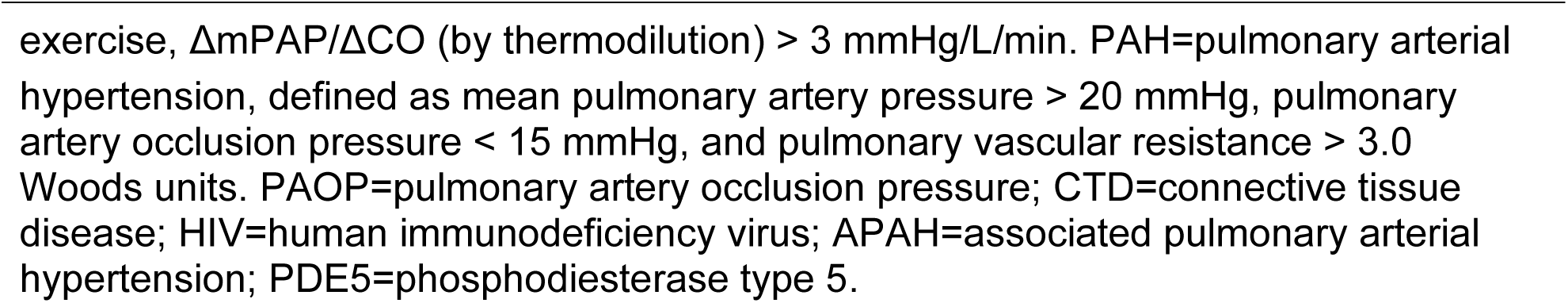
Baseline characteristics of subjects, by endothelial cell samples.

|  | **PAH** | **Controls** |
| --- | --- | --- |
| Balloon tips* | 32 | 3 |
| Subjects | 25 | 3 |
| Female sex | 29 (91) | 3 (100) |
| Age, yr | 56 (30 – 84) | 63 (19 – 78) |
| Body mass index, kg/m<sup>2</sup> | 29 (16 – 45) | 22 (21 – 31) |
| Self-reported race |  |  |
| White | 26 (81) | 2 (67) |
| Black | 2 (6) | . |
| Asian | 1 (3) | . |
| Other | 3 (9) | 1 (33) |
| Self-reported Hispanic ethnicity | 4 (13) | 1 (33) |
| Hemodynamics |  |  |
| Right atrial pressure, mm Hg | 8 (3 – 19) | 7 (1 – 7) |
| Mean pulmonary artery pressure, mm Hg | 39 (16 – 69) | 18 (16 – 21) |
| PAOP, mm Hg | 11 (5 – 17) | 10 (8 – 12) |
| Cardiac output, L/min | 5.5 (1.9 – 12.9) | 5.3 (4.9 – 5.6) |
| Pulmonary vascular resistance, Wood units | 4.0 (1.0 – 19.6) | 1.8 (1.1 – 1.9) |
| PAH etiology |  |  |
| Idiopathic PAH | 8 (25) | . |
| Heritable PAH | 3 (9) | . |
| CTD-APAH | 9 (28) | . |
| Congenital heart disease-APAH | 2 (6) | . |
| HIV-APAH | 1 (3) | . |
| Portopulmonary hypertension | 5 (16) | . |
| Pulmonary veno-occlusive disease | 2 (6) | . |
| Exercise-induced pulmonary hypertension | 2(6) | . |
| Functional class III/IV, *n*=7 | 2 (29) | . |
| Six-minute walk distance, m, *n*=5 | 375 (150 – 436) | . |
| Brain natriuretic level, pg/mL, *n*=5 | 359 (34 – 1141) | . |
| REVEAL 2.0 Risk Score, *n*=28 | 10 (4 – 12) | . |
| PAH treatments |  | . |
| None | 2 (6) | . |
| Calcium channel blockers | 0 | . |
| PDE5 inhibitors | 24 (75) | . |
| Endothelial receptor antagonists | 4 (13) | . |
| Prostacyclin pathway | 2 (6) | . |
| Diuretics, *n*=25 | 20 (80) | . |
| Psychotropics, Serotonin | 8 (32) | . |
| Psychotropics, Non-Serotonin | 7 (28) | . |

Data represented as n (%) or median (range). \*25 subjects had a total of 7 biological replicates (1 subject with 5 balloon tips, 3 subjects with 2 balloon tips). Exercise-induced pulmonary hypertension defined as resting mPAP < 20 mmHg and with
exercise, $\Delta mPAP/\Delta CO$ (by thermodilution) $> 3$ mmHg/L/min. PAH=pulmonary arterial hypertension, defined as mean pulmonary artery pressure $> 20$ mmHg, pulmonary artery occlusion pressure $< 15$ mmHg, and pulmonary vascular resistance $> 3.0$ Woods units. PAOP=pulmonary artery occlusion pressure; CTD=connective tissue disease; HIV=human immunodeficiency virus; APAH=associated pulmonary arterial hypertension; PDE5=phosphodiesterase type 5.

### Transcriptomic Profile of Participants

The PAEC RNA-Seq transcriptomic dataset, after filtering, included 32 PAH samples (from 25 unique PAH patients), 3 non-disease control samples, and 12,562 genes. The multidimensional scaling plot of the dataset shows that the variance in gene expression explained by the first and second principal component vectors are 30% and 9%, respectively, with separation between the non-disease controls and PAH groups, but also substantial separation within the PAH group (**Figure S1A**). The coefficient of variation (%CV) range for the expression of genes among PAH samples was 33–545%, with 6,545 (52%) genes having a %CV ≥80% (**Figure S1B**). The average correlation coefficient for replicates in non-disease controls is 0.91, whereas the average correlation coefficient of replicates in PAH samples is 0.81 (**Figure S2**). These collective data suggest important pathobiological variability across the PAH samples and are consistent with the clinical heterogeneity observed in this (and many other) study populations^1, 25, 26^.

### PAH Endothelial Cell Biopsy-Derived Individualized PPI Networks

The individualized PAH PPI networks varied widely in topology (median=5954 [2074 min, 9569 max] number of nodes) and complexity (median=8494 [2097 min, 24877 max] number of edges) (**Table 2, Figure S3**). The expression pattern of PAH molecular targets across individualized networks is presented in **Figure S4**. We used the overlap coefficient, a similarity measure that measures the overlap between two finite sets, to evaluate the node similarity and edge similarity of two individualized PAH networks^13^. The pair-wise node similarity of individualized networks displays a modular pattern, with an average 0.77. The edge similarity of individualized networks varies substantially, from 0.12 to 0.57 (**Figure S5**). A much greater overlap for nodes was observed compared with edges across the PAH networks. The uniqueness of sample-specific networks (i.e., the nodes and edges that do not appear in any other networks) varied from 0 to 37 nodes (median=3), and from 8 to 4119 edges (median=219). This finding emphasizes that it is the wiring diagrams (interactions), rather than the proteins themselves that distinguished patients in this study. Topological analyses of the individualized PAH PPI networks show that, while all the networks have similar density, they have a wide range of network diameter, characteristic path length, and size of largest connected components (**Table S4**).

**Table 2.**
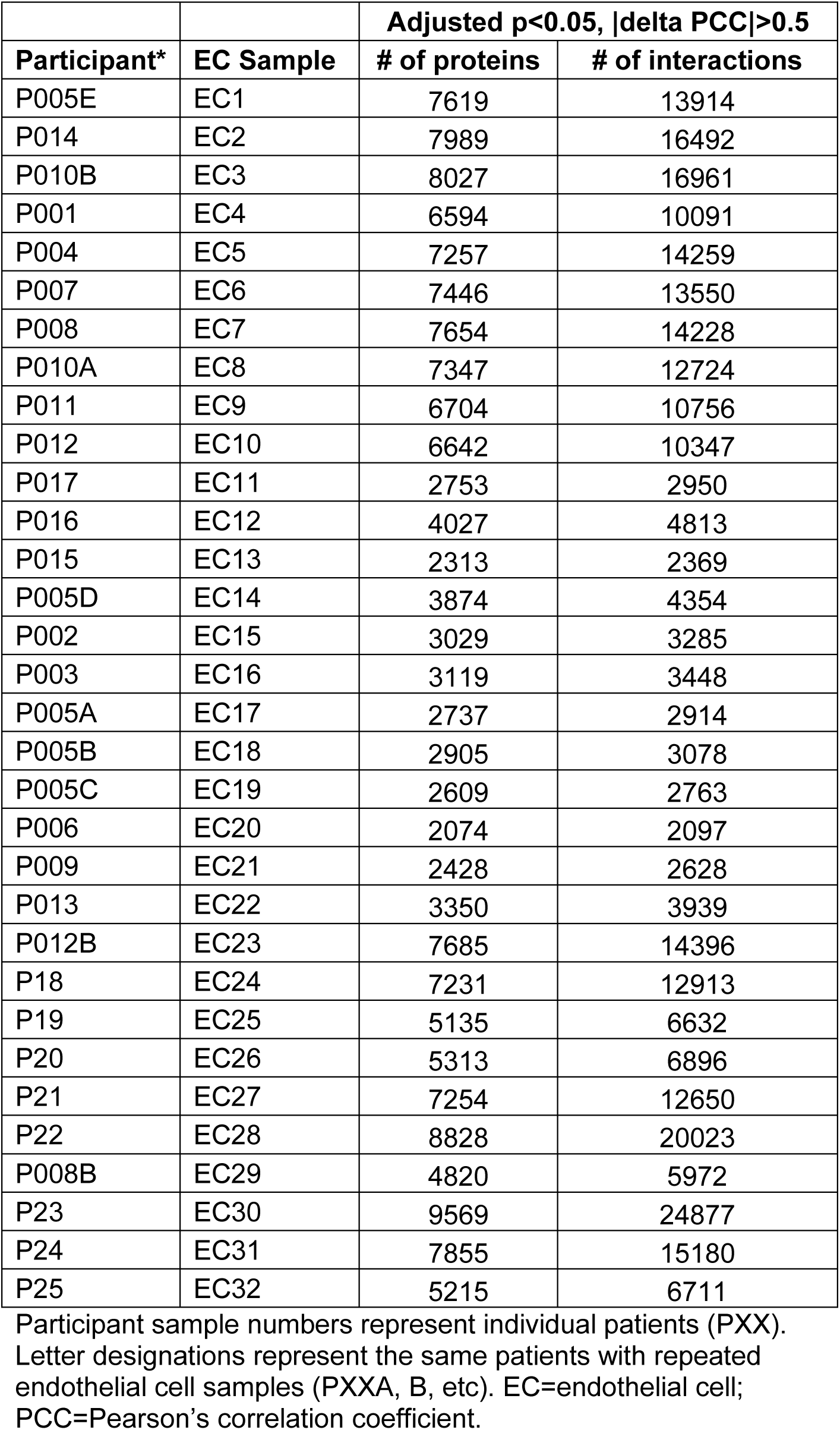
Protein-protein Interaction Network details, by endothelial cell sample and after adjustment for multiple comparisons.

We next performed a subgroup-level analysis to determine if unique molecular signatures aligned with clinical phenotypes. Based on the multidimensional scaling and edge similarity data, three distinct subclusters were identified by using hierarchical clustering (**Table S5, Figure S6**), reinforcing the utility of the network-based method for segregating distinct PPI among patients that share the same diagnosis of PAH. Moreover, across these clusters there were no clear clinical phenotypic differences.

The principal clinical features of PAH include a variety of phenotypes, such as fibrosis, inflammation, levels of oxidative stress, hypertrophy, etc^27^. To demonstrate that our patient-specific networks can capture individual-level pathobiological differences and phenotypic heterogeneity, we conducted an endophenotype enrichment analysis. Out of 75 endophenotypes we compiled, 34 endophenotypes were significantly (p < 0.05 adjusted by the Benjamini-Hochberg procedure) enriched in at least one sample. A heatmap of -log_10_P is shown in **Figure S7**. All of the individualized PAH interactomes were significantly enriched with genes associated with hypoxia, oxidative stress, apoptosis and the majority of the individualized PAH interactomes were significantly enriched with DNA damage, angiogenesis, cell proliferation and immune response. There was greater variation across PAH patients in enrichment for genes associated with fibrosis, thrombosis, and inflammation. The results demonstrate the phenotypic heterogeneity of these patients and the capability of patient-specific networks to capture individual-level differences.

### PAH therapeutic target-interactome alignment and PAH endpoints

We identified a PAH, diuretic, serotonin psychotropic and non-serotonin psychotropic pharmacotherapeutic target in 31, 3, 9 and 2 individualized interactomes, respectively. A heatmap of proportion of targets within a drug class expressed by each participant is provide in **Figure 2A**. The quantitative rank-order summary of patient-specific PAH and comparator pharmacotherapeutic molecular targets is provided in **Tables S6-10**, which indicates a non-normal distribution of targets in individualized networks for either PAH or comparator drugs. Notably, there was wide variation in the proportion of individuals expressing targets for PAH-specific classes (**Figure 2B**).

**Figure 2.**
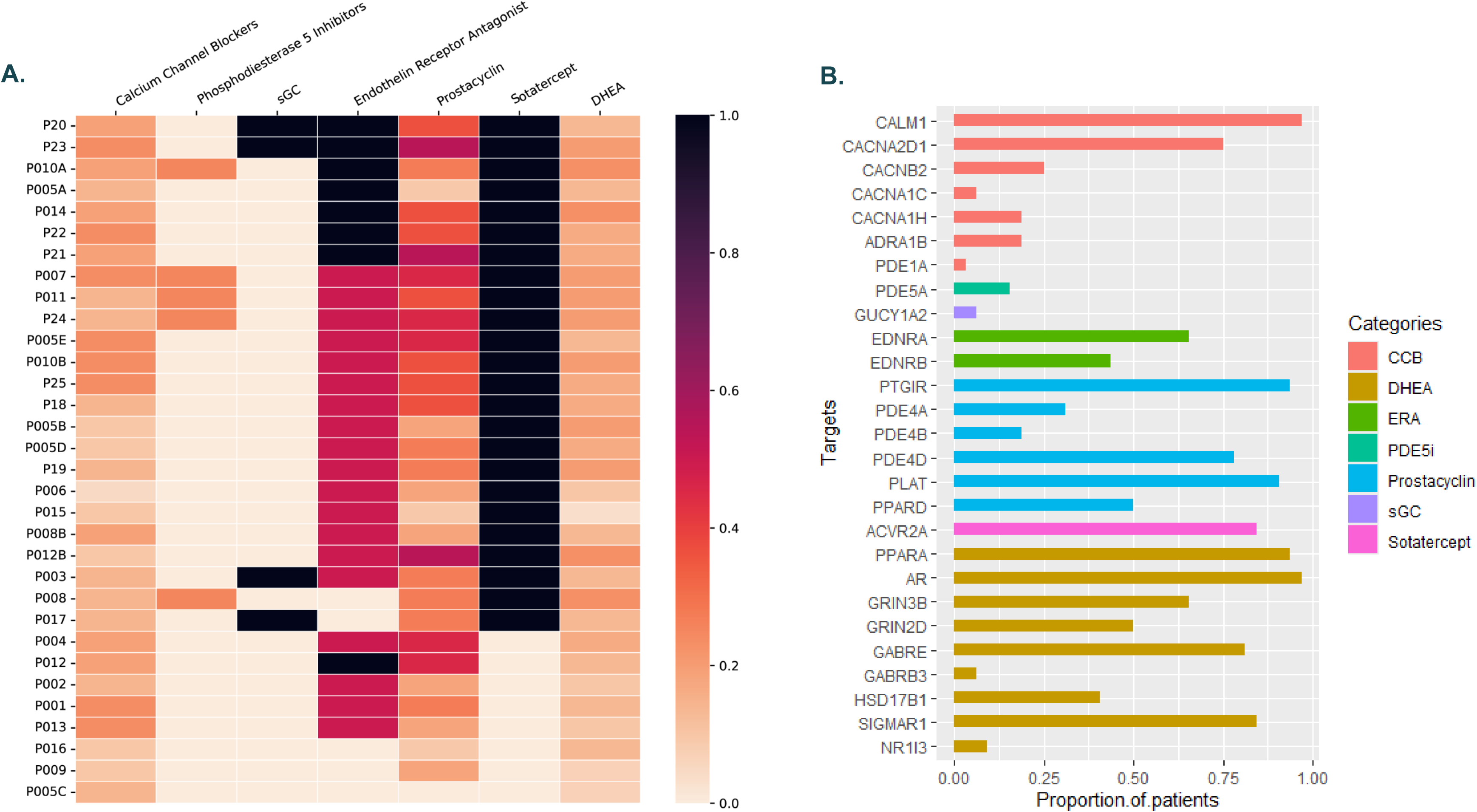
Heterogeneity of patient-specific PAH pharmacotherapeutic molecular targets expressed in cell biopsy samples. **A)** Heatmap of proportion of targets within a drug class expressed by each participant. Row=individual participant. Column=drug class. Cell=proportion of targets expressed by an individual patient. Purple=100% targets expressed. Peach=0% targets expressed. sGC=stimulator of soluble guanylate cyclase. DHEA=dehydroepiandrosterone. **B)** Proportion of participants who express each target. CCB=calcium channel blocker. PDE5i=phosphodiesterase 5 inhibitor. sGC=stimulator of soluble guanylate cyclase. ERA=endothelin receptor antagonist. DHEA=dehydroepiandrosterone.

Greater within-class therapeutic target-interactome alignment was associated with improvements in multiple PAH end points. For example, complete concordance was associated with over a ∼160 m increase at 6 months and a ∼55 m increase in 6MWD at 12 months, although the overall associations did not reach statistical significance likely due to sample size (e.g., 6 month β 168 meters [95% confidence interval (CI) 106.1, 442.3], p = 0.20)) (**Figures 3A and B**). Similarly, we observed a transition in BNP levels from elevated at baseline to within the normal range that corresponded to an absolute decrease of ∼627 pg/mL (β -627.6, 95% CI -986.4, - 268.8]; p = 0.003) at 6 months (**Figure 3C**), an association which persisted but was not significant at 12 months (β -129.6, 95% CI -338.8, 79.6]; p = 0.22) (**Figure 3D**). Therapeutic class target-interactome alignment was also likely associated with WHO FC improvement at 6 (p = 0.11) and 12 months (p = 0.06) (**Figures 3E and F**).

**Figure 3.**
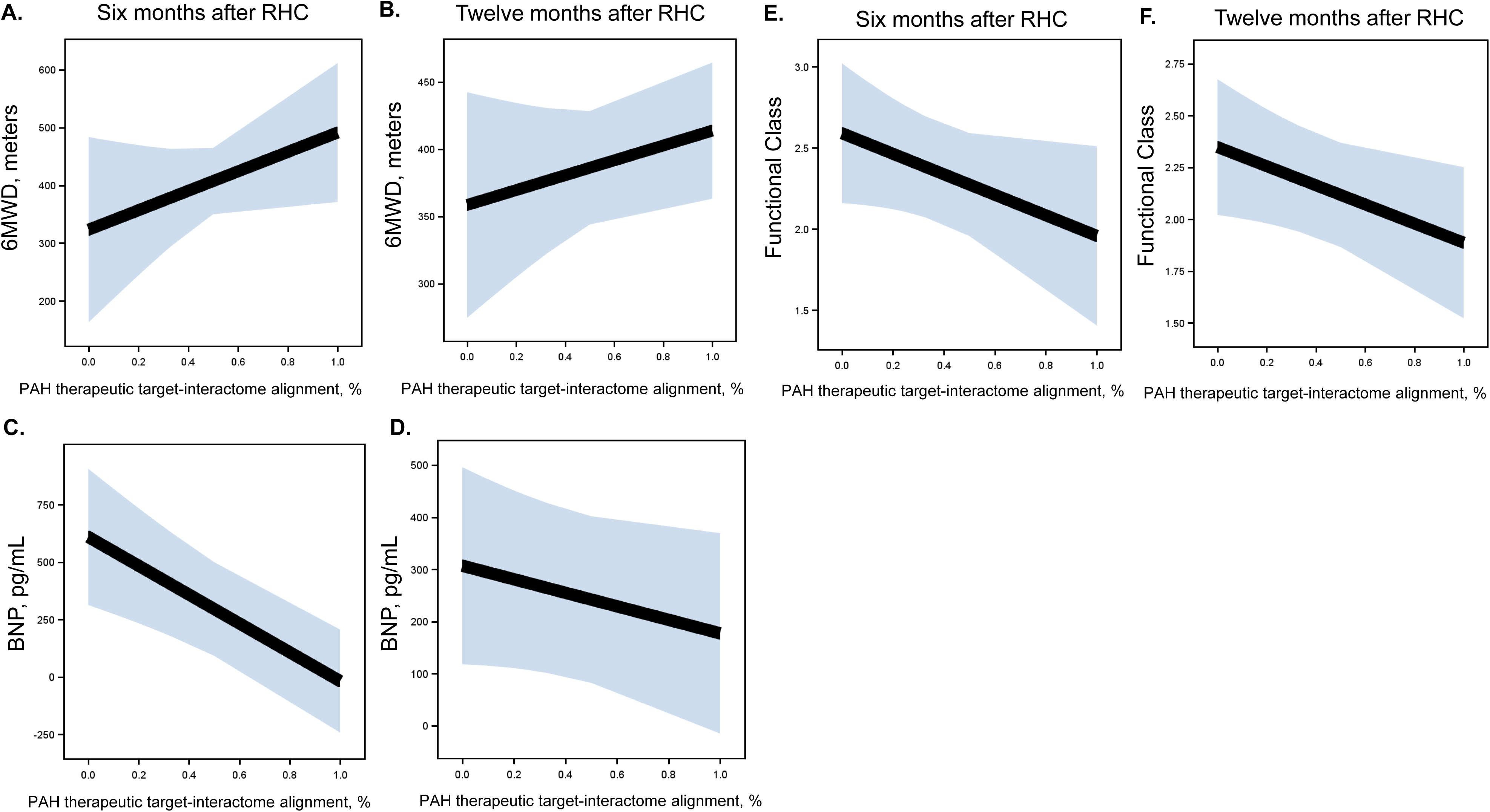
Association between PAH therapeutic target-interactome alignment and six-minute walk distance (6MWD) (**Panels A, B**), brain natriuretic peptide (BNP) levels (**Panels C, D**) and functional class (**Panels E, F**) at six and twelve months after right heart catheterization (RHC).

Greater therapeutic target-interactome alignment was associated with a significant improvement in REVEAL 2.0 Risk score at 6 months (β -1.3, [95% CI -2.6, -0.04]; p=0.04) (**Figure 4A**) and 12 months (β -1.7, [95% CI -3.4, 0.07]; p=0.06) (**Figure 4B**), generally ranging from intermediate (7 – 8) to high risk (≥ 9) for no alignment to intermediate-low risk (≤ 8) for mortality with complete alignment. There may have also been an association between greater therapeutic class target-treatment concordance and lower COMPERA 2.0 four-strata risk assessments at 6 months (β -2.5 [95% CI -16.4, 11.3]; p = 0.26) (**Figure 4C**), a relationship not observed at 12 months (**Figure 4D**).

**Figure 4.**
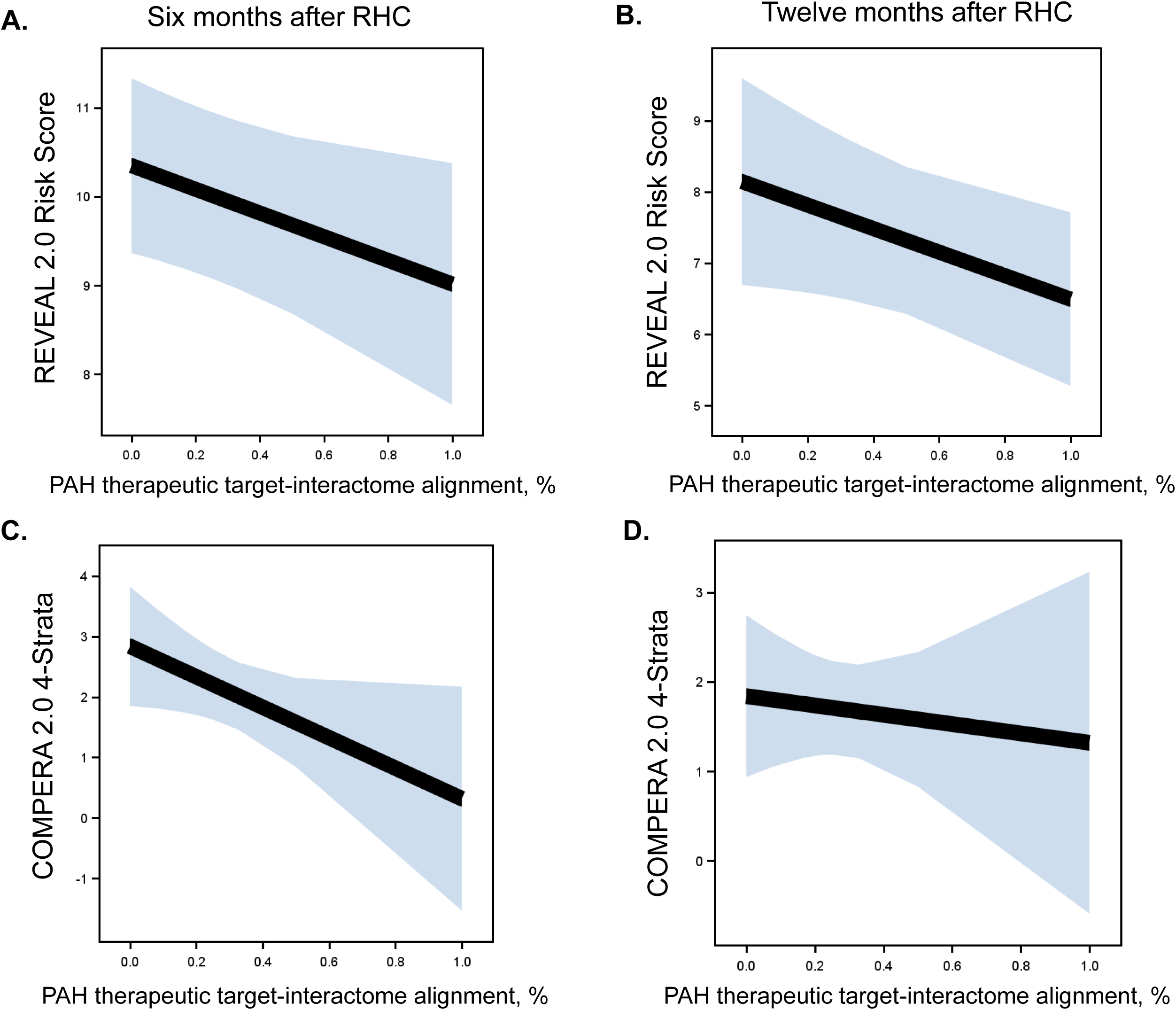
Association between PAH therapeutic target-interactome alignment and improvement in the United States Registry to Evaluate Early and Long-Term PAH Disease Management (REVEAL) 2.0 risk score (**Panels A, B**) and the Comparative, Prospective Registry of Newly Initiated Therapies for PH (COMPERA) 2.0 four-strata risk assessment (**Panels C, D**) at six and twelve months after right heart catheterization (RHC), respectively.

### Comparator target-interactome alignment and PAH endpoints

Among the 32 participants in this study, 20 (80%), 8 (32%) and 7 (28%) were prescribed diuretics, or drugs included in the Serotonin Psychotropic and Non-Serotonin Psychotropic groups, respectively, at the time of RHC. Spironolactone was the only diuretic for which ≥1 drug-specific molecular target(s) were found to overlap in the PAH module but was prescribed too infrequently (n=1) in this study population for further analysis. Among participants prescribed diuretics exclusive of spironolactone at the time of RHC, there was no relationship identified between therapeutic target-interactome alignment and 6MWD, BNP levels, or WHO-FC at 6- or 12-months (data not shown). Similarly, there were no associations identified between diuretic target-interactome alignment and risk scores at 6- or 12-months (data not shown).

Among those prescribed Serotonin Psychotropic drugs, increasing alignment was associated with worsening REVEAL 2.0 scores. As concordance increased, REVEAL 2.0 Risk scores worsened from low to high risk for mortality at 6 months (p = 0.03), a relationship which was attenuated at 12 months (p = 0.18) (**Figure S8**). Conversely, participants that were prescribed Non-Serotonin Psychotropics at the time of RHC and had target alignment were significantly more likely to have lower REVEAL 2.0 Risk scores at 6 months, such that alignment in gene targets was associated with a change from high risk (≥9) to intermediate risk (7-8) (p = 0.050), a relationship which was not observed at 12 months (p = 0.59). Gene target-Non-Serotonin Psychotropic alignment had no detectable relationship with other PAH end points.

## Discussion

In this study, a novel network medicine approach integrating transcriptomics from PAEC biopsies acquired from PAH patients at point-of-care was used to link individualized treatment selection with improvement in clinically relevant end points. In doing so, this work progresses precision medicine through three key advances. First, we established an analytical pipeline integrating functional PPIs, derived from patient cells *ex vivo* and modeled *in silico,* with systems pharmacology to anticipate patient-level therapeutic responses. Second, we demonstrated the potential for individualized interactomes to harbor clinically actionable data, which we tested with a novel study design pairing patient-specific pathobiological readouts to drug effects. Third, findings from this study emphasize the broad translational potential of cell biopsies collected during routine clinical practice despite (or perhaps as a function of) variability across samples. Taken together, these findings provide a basis for further prospective studies scaling patient-specific treatment regimens based on individualized therapeutic response to a standard-of-care approach in PAH, with implications for other complex diseases characterized by both diverse pathogenetic causes and variable outcomes.

Greater appreciation of the clinical and molecular diversity underpinning pulmonary vascular diseases has stimulated efforts to reclassify PAH patients based on deep phenotyping. Distinct endophenotypes determined by peripheral blood signatures (i.e., inflammation, monogenic variants, others) are associated with differences in PAH severity and outcome^25, 28–33^. However, approaches used in these prior studies tended to use analytic methods that may overemphasize single molecular events or bystander (rather than functional) biomarkers to explain pathogenicity and have not been individualized or utilized to inform treatment selection successfully. Here, we used (valid) functional PPIs to assemble biopsy-specific interactomes, characterize pathobiological patterns between patients, and test PAH pharmacotherapeutic response. We observed general agreement in network topology across patients when considering proteins (nodes), but substantial variability ranging from near-total to near-absent overlap between patients (including in repeated samples from the same patients) when focusing on functional PPIs. These findings imply that phenotypic heterogeneity across patients (and disease course) with PAH hinges on molecular connectivity detectable in affected tissue (i.e., PAECs), rather than simple expression profiles. This has important implications for treatment in PAH, since uncoupling between transcript quantity and pathogenicity is common in complex diseases^34^, whereas PPIs implicated in individualized interactome connectivity itself appeared in this study to be a measure of functional importance.

Despite acknowledgement of the need for deep phenotyping and precision-based treatment approaches in PAH, especially given the evolving treatment landscape, challenges remain in operationalizing these concepts. Because PAH remains a highly morbid disease, guidelines have understandably emphasized initial combination therapy and treatment escalation to improve outcomes^5^. Treatment response heterogeneity has been identified in PAH clinical trials and registries based on patient comorbidities^2, 3^ which could inform prognostic enrichment, but an equally important goal would be to enhance drug development and clinical trial efficiency via predictive enrichment (e.g., targeting patient-level molecular signatures). At the bedside, clinician selection of precise PAH therapeutic regimens would ideally improve outcomes and health-related quality of life while minimizing side effects, burden, and cost. This proof-of-concept *in silico* trial – which derives individualized networks from cell biopsies of the affected tissue compartment paired to prescribed treatment and important outcomes in PAH – is the first step towards a point-of-care process for therapeutic efficacy and effectiveness screening and personalized medicine in PAH. Optimizing the analytic pipeline by scaling the cell biopsy approach, implementing real-time transcriptomics, and semi-automating the bioinformatic pipeline is necessary to validate the applicability and feasibility of the methods.

Gene target-PAH therapeutic class alignment was associated with improvement in multiple important PAH end points used for drug approval or treatment escalation in clinical guidelines. Some of these relationships persisted for up to a year after introduction of therapy. The effect estimate observed for 6MWD at 6 months – over 160 m – well surpassed the minimally clinically important difference of approximately 33 m for 6MWD in PAH^15^. Gene target-PAH therapeutic class alignment was also associated with normalization of BNP levels to low categories of PAH risk^5^. Estimates for REVEAL 2.0 Risk and COMPERA 2.0 risk assessments were decreased by at least one risk category, which has been associated with improved survival and a reduction in important clinical events including lung transplant and hospitalizations^16, 35^. These signals were detected despite a relatively small sample size, the non-controlled study design and background therapies, data missingness (addressed below), and analyses at the therapeutic class (not specific drug) level, which would all serve to increase the signal to noise ratio and bias our results to the null.

Serotonin synthesis and signaling is implicated in PAH pathobiology^22, 24^. Many observational studies have demonstrated relationships between antidepressants, specifically selective serotonin reuptake inhibitors, and risk in variable forms of pulmonary vascular disease^23^. More recently, rodatristat ethyl was developed to enhance serotonin synthesis by peripheral inhibition of tryptophan hydroxylase 1 (TPH1) and tested in a Phase 2b study terminated for safety^36^. We observed similar relationships in the current study, where psychotropic medications targeting serotonin signaling were possibly associated with worse PAH metrics. Confirmation of overlap of these drugs in the PAH module, detection of related gene targets in our cell biopsy samples, and linking of greater target engagement with PAH outcomes in a direction that was predicted by empiric research provides key measure of validity of to our approach as well as insight into molecular phenotypes in PAH patients *per se* that may be informative in clinical studies involving these drugs.

There were no relationships detected between PPI alignment and diuretics outside of the PAH module (a pure “placebo” condition) and PAH end points, as hypothesized given no direct molecular targets were found (nor anticipated) in the PAH module. Target alignment with Non-Serotonin Psychotropics were associated with improved REVEAL 2.0 Risk scores, but had no associations with 6MWD, BNP or functional class. As these treatments had no overlap in the PAH module (and therefore would be unlikely to target biologic pathways relevant to pulmonary vascular disease in the study participants), we speculate this could be due to confounding by indication. In other words, patients who were experiencing progressive disease (i.e., high REVEAL 2.0 Risk scores at the time of cell biopsy) were more likely to be prescribed psychotropic medications, given the known high concurrent rate of anxiety/depression with PAH^37, 38^, with subsequent reduction in REVEAL scores over time with PAH treatment.

Although findings from this study advance personalized therapeutic selection using clinically relevant evidence, prospective research with protocolized study design and clinical assessments (e.g., a randomized clinical trial) is needed to clarify the translatability of our methodology (and derivative findings) to clinical practice. These samples were primarily collected over the course of the COVID-19 pandemic and as such there was more missing clinical data after treatments were introduced than would be expected, and we cannot fully know the mechanisms of missingness (at random, not at random)^39^. The overall study population size was modest and the size of some subgroups (e.g., patients prescribed spironolactone) limited statistical analyses. Males were underrepresented in this study and, therefore, sex imbalance is an important potential confounder of our results. In the case of REVEAL 2.0 Risk scores some missing values were assigned a value of “0” albeit using a validated approach^16^. As we do not routinely perform RHC at standard intervals for all patients clinically, selection or referral bias may have influenced our results. We did not routinely collect data on HRQoL or therapeutic burden, although this will be important in future precision-based efforts.

## Conclusion

Pulmonary endothelial biopsy data collected at point-of-care in PAH and analyzed using a network medicine-systems pharmacology pipeline harbors clinically actionable information for PAH patients. Findings from this study advance precision medicine into a framework in which individualized clinical decision-making is possible and warrants future prospective *in silico* studies validating bioinformatics-based approaches to treatment selection and clinical trial design in PAH, as well as other complex diseases defined by pathophenotypic heterogeneity.

## Data Availability

If accepted, all code and relevant data will be deposited into publicly available repository in accordance with Journal policies.

## Acknowledgements

The authors would like to acknowledge all the patients who participated in these studies.

## Author Contributions

B.A.M. and C.E.V. project conception, study design, study planning, partial funding, data interpretation, partial data interpretation, manuscript drafting, manuscript editing; R.S.W., N.S. study design and planning, network derivation, data analysis, manuscript editing; G.B. data analysis and interpretation; H.W., K.C.F., J.R.K., C.J.M., M.W. data collection and participant recruitment, revision of the manuscript; M.P., E.O.H. cell biopsy method, manuscript revision.

## References

1. Pan HM, McClelland RL, Moutchia J, et al. Heterogeneity of treatment effects by risk in pulmonary arterial hypertension. Eur Respir J 2023; 62 20230727. DOI: 10.1183/13993003.00190-2023.

2. Moutchia J, McClelland RL, Al-Naamani N, et al. Pulmonary arterial hypertension treatment: an individual participant data network meta-analysis. Eur Heart J 2024; 45: 1937–1952. DOI: 10.1093/eurheartj/ehae049.

3. Hoeper MM, Dwivedi K, Pausch C, et al. Phenotyping of idiopathic pulmonary arterial hypertension: a registry analysis. Lancet Respir Med 2022; 10: 937–948. 20220628. DOI: 10.1016/s2213-2600(22)00097-2.

4. Hoeper MM, Pausch C, Grünig E, et al. Idiopathic pulmonary arterial hypertension phenotypes determined by cluster analysis from the COMPERA registry. J Heart Lung Transplant 2020; 39: 1435–1444. 20200930. DOI: 10.1016/j.healun.2020.09.011.

5. Humbert M, Kovacs G, Hoeper MM, et al. 2022 ESC/ERS Guidelines for the diagnosis and treatment of pulmonary hypertension: Developed by the task force for the diagnosis and treatment of pulmonary hypertension of the European Society of Cardiology (ESC) and the European Respiratory Society (ERS). Endorsed by the International Society for Heart and Lung Transplantation (ISHLT) and the European Reference Network on rare respiratory diseases (ERN-LUNG). Eur Heart J 2022; 43: 3618–3731. DOI: 10.1093/eurheartj/ehac237.

6. Budhiraja R, Tuder RM and Hassoun PM. Endothelial dysfunction in pulmonary hypertension. Circulation 2004; 109: 159–165. 2004/01/22. DOI: 10.1161/01.cir.0000102381.57477.50.

7. Evans CE, Cober ND, Dai Z, et al. Endothelial cells in the pathogenesis of pulmonary arterial hypertension. Eur Respir J 2021; 58 20210902. DOI: 10.1183/13993003.03957-2020.

8. Ryanto GRT, Ikeda K, Miyagawa K, et al. An endothelial activin A-bone morphogenetic protein receptor type 2 link is overdriven in pulmonary hypertension. Nat Commun 2021; 12: 1720. DOI: 10.1038/s41467-021-21961-3.

9. Ventetuolo CE, Aliotta JM, Braza J, et al. Culture of pulmonary arterial endothelial cells from pulmonary artery catheter balloon tips: Considerations for use in pulmonary vascular disease. Eur Respir J 2020 2020/01/18. DOI: 10.1183/13993003.01313-2019.

10. Singh N, Eickhoff C, Garcia-Agundez A, et al. Transcriptional profiles of pulmonary artery endothelial cells in pulmonary hypertension. Sci Rep 2023; 13: 22534. 20231218. DOI: 10.1038/s41598-023-48077-6.

11. Pollett JB, Benza RL, Murali S, et al. Harvest of pulmonary artery endothelial cells from patients undergoing right heart catheterization. J Heart Lung Transplant 2013; 32: 746–749. 2013/05/21. DOI: 10.1016/j.healun.2013.04.013.

12. Tielemans B, Stoian L, Wagenaar A, et al. Incremental Experience in In Vitro Primary Culture of Human Pulmonary Arterial Endothelial Cells Harvested from Swan-Ganz Pulmonary Arterial Catheters. Cells 2021; 10 20211119. DOI: 10.3390/cells10113229.

13. Maron BA, Wang RS, Shevtsov S, et al. Individualized interactomes for network-based precision medicine in hypertrophic cardiomyopathy with implications for other clinical pathophenotypes. Nat Commun 2021; 12: 873. 20210208. DOI: 10.1038/s41467-021-21146-y.

14. Simonneau G, Gatzoulis MA, Adatia I, et al. Updated clinical classification of pulmonary hypertension. J Am Coll Cardiol 2013; 62: 029. Research Support, NIH , Extramural Research Support, Non-U S Gov’t Review.

15. Moutchia J, McClelland RL, Al-Naamani N, et al. Minimal clinically important difference in the 6-minute-walk distance for patients with pulmonary arterial hypertension. Am J Respir Crit Care Med 2023; 207: 1070–1079. DOI: 10.1164/rccm.202208-1547OC.

16. Benza RL, Gomberg-Maitland M, Elliott CG, et al. Predicting survival in patients with pulmonary arterial hypertension: The REVEAL Risk Score Calculator 2.0 and comparison with ESC/ERS-based risk assessment strategies. Chest 2019; 156: 323–337. 2019/02/18. DOI: 10.1016/j.chest.2019.02.004.

17. Boucly A, Weatherald J, Savale L, et al. External validation of a refined four-stratum risk assessment score from the French pulmonary hypertension registry. Euro Respir J 2022; 59 60: 2102419. DOI: 10.1183/13993003.02419-2021.

18. Walsh TP, Baird GL, Atalay MK, et al. Experimental design of the Effects of Dehydroepiandrosterone in Pulmonary Hypertension (EDIPHY) trial. Pulm Circ 2021; 11: 2045894021989554. 2021/06/08. DOI: 10.1177/2045894021989554.

19. Hoeper MM, Badesch DB, Ghofrani HA, et al. Phase 3 trial of sotatercept for treatment of pulmonary arterial hypertension. New Engl J Med 2023; 388: 1478–1490. DOI: 10.1056/NEJMoa2213558.

20. Humbert M, McLaughlin V, Gibbs JSR, et al. Sotatercept for the treatment of pulmonary arterial hypertension. New Eng J Med 2021; 384: 1204–1215. DOI: 10.1056/NEJMoa2024277.

21. Wang RS and Loscalzo J. Network module-based drug repositioning for pulmonary arterial hypertension. CPT Pharmacometrics Syst Pharmacol 2021; 10: 994–1005. 20210709. DOI: 10.1002/psp4.12670.

22. Naeije R and Eddahibi S. Serotonin in pulmonary arterial hypertension. Am J Respir Crit Care Med 2004; 170: 209–210. DOI: 10.1164/rccm.2406002.

23. Sadoughi A, Roberts KE, Preston IR, et al. Use of selective serotonin reuptake inhibitors and outcomes in pulmonary arterial hypertension. Chest 2013; 144: 531–541. DOI: 10.1378/chest.12-2081.

24. Maron BA. Inhibiting serotonin to treat pulmonary arterial hypertension. JACC: Basic Transl Sci 2023; 8: 1389–1391. DOI: doi:10.1016/j.jacbts.2023.09.008.

25. Kariotis S, Jammeh E, Swietlik EM, et al. Biological heterogeneity in idiopathic pulmonary arterial hypertension identified through unsupervised transcriptomic profiling of whole blood. Nat Commun 2021; 12: 7104. 20211207. DOI: 10.1038/s41467-021-27326-0.

26. Chelladurai P, Savai R and Pullamsetti SS. Zooming into cellular and molecular heterogeneity of pulmonary hypertension. what more single-cell omics can offer. Am J Respir Crit Care Med 2021; 203: 941–943. DOI: 10.1164/rccm.202010-3889ED.

27. Dweik RA, Rounds S, Erzurum SC, et al. An Official American Thoracic Society Statement: Pulmonary Hypertension Phenotypes. Am J Respir Crit Care Med 2014; 189: 345–355. DOI: 10.1164/rccm.201311-1954ST.

28. Rhodes CJ, Ghataorhe P, Wharton J, et al. Plasma metabolomics implicates modified transfer rnas and altered bioenergetics in the outcomes of pulmonary arterial hypertension. Circulation 2017; 135: 460–475. 2016/11/25. DOI: 10.1161/circulationaha.116.024602.

29. Rhodes CJ, Otero-Núñez P, Wharton J, et al. Whole-blood RNA profiles associated with pulmonary arterial hypertension and clinical outcome. Am J Respir Crit Care Med 2020; 202: 586–594. DOI: 10.1164/rccm.202003-0510OC.

30. Harbaum L, Rhodes CJ, Wharton J, et al. Mining the plasma proteome for insights into the molecular pathology of pulmonary arterial hypertension. Am J Respir Crit Care Med 2022; 205: 1449–1460. DOI: 10.1164/rccm.202109-2106OC.

31. Soon E, Holmes AM, Treacy CM, et al. Elevated levels of inflammatory cytokines predict survival in idiopathic and familial pulmonary arterial hypertension. Circulation 2010; 122: 920–927. 2010/08/18. DOI: 10.1161/circulationaha.109.933762.

32. Sweatt AJ, Hedlin HK, Balasubramanian V, et al. Discovery of distinct immune phenotypes using machine learning in pulmonary arterial hypertension. Circ Res 2019; 124: 904–919. 2019/01/22. DOI: 10.1161/circresaha.118.313911.

33. Rhodes CJ, Batai K, Bleda M, et al. Genetic determinants of risk in pulmonary arterial hypertension: international genome-wide association studies and meta-analysis. Lancet Respir Med 2019; 7: 227–238. 20181205. DOI: 10.1016/s2213-2600(18)30409-0.

34. Vidal M, Cusick ME and Barabási AL. Interactome networks and human disease. Cell 2011; 144: 986–998. DOI: 10.1016/j.cell.2011.02.016.

35. Min J, Badesch D, Chakinala M, et al. Prediction of health-related quality of life and hospitalization in pulmonary arterial hypertension: The Pulmonary Hypertension Association Registry. Am J Respir Crit Care Med 2021; 203: 761–764. DOI: 10.1164/rccm.202010-3967LE.

36. Lazarus HM, Denning J, Wring S, et al. A trial design to maximize knowledge of the effects of rodatristat ethyl in the treatment of pulmonary arterial hypertension (ELEVATE 2). Pulm Circ 2022; 12: e12088. 20220511. DOI: 10.1002/pul2.12088.

37. Harzheim D, Klose H, Pinado FP, et al. Anxiety and depression disorders in patients with pulmonary arterial hypertension and chronic thromboembolic pulmonary hypertension. Respir Res 2013; 14: 104. 20131009. DOI: 10.1186/1465-9921-14-104.

38. McCollister DH, Beutz M, McLaughlin V, et al. Depressive symptoms in pulmonary arterial hypertension: prevalence and association with functional status. Psychosomatics 2010; 51: 339–339.e338. DOI: 10.1176/appi.psy.51.4.339.

39. Hossie TJ, Gobin J and Murray DL. Confronting missing ecological data in the age of pandemic lockdown. Front Ecol Evol 2021.

